# A Molecular network approach reveals shared cellular and molecular signatures between chronic fatigue syndrome and other fatiguing illnesses

**DOI:** 10.1101/2021.01.29.21250755

**Authors:** Phillip H. Comella, Edgar Gonzalez-Kozlova, Roman Kosoy, Alexander W. Charney, Irene Font Peradejordi, Shreya Chandrasekar, Scott R. Tyler, Wenhui Wang, Bojan Losic, Jun Zhu, Gabriel E. Hoffman, Seunghee Kim-Schulze, Jingjing Qi, Manishkumar Patel, Andrew Kasarskis, Mayte Suarez-Farinas, Zeynep H. Gümüş, Carmen Argmann, Miriam Merad, Christian Becker, Noam D. Beckmann, Eric E. Schadt

## Abstract

The molecular mechanisms of chronic fatigue syndrome (CFS, or Myalgic encephalomyelitis), a disease defined by extreme, long-term fatigue, remain largely uncharacterized, and presently no molecular diagnostic test and no specific treatments exist to diagnose and treat CFS patients. While CFS has historically had an estimated prevalence of 0.1-0.5% [1], concerns of a “long hauler” version of Coronavirus disease 2019 (COVID-19) that symptomatically overlaps CFS to a significant degree **(Supplemental Table-1)** and appears to occur in 10% of COVID-19 patients[2], has raised concerns of a larger spike in CFS [3]. Here, we established molecular signatures of CFS and a corresponding network-based disease context from RNA-sequencing data generated on whole blood and FACs sorted specific peripheral blood mononuclear cells (PBMCs) isolated from CFS cases and non-CFS controls. The immune cell type specific molecular signatures of CFS we identified, overlapped molecular signatures from other fatiguing illnesses, demonstrating a common molecular etiology. Further, after constructing a probabilistic causal model of the CFS gene expression data, we identified master regulator genes modulating network states associated with CFS, suggesting potential therapeutic targets for CFS.

## Main Text

Considerable controversy exists as to whether CFS has one or many causes [4, 5] and whether the resulting symptoms are somatic or psychosomatic [6]. Previous studies have proposed viral infections as a possible cause of CFS [7, 8], but to date, no single (or set) of virus(es) has been reported to be causally associated to the syndrome. Other hypotheses have been brought forth, such as immune system abnormalities[9], NK and T cell dysfunction[10, 11], cytokine dysregulations [12, 13], endocrinologic and metabolic abnormalities[14, 15], and miRNA associations with severity[16]. With the lack of understanding of CFS, there is a scarcity of potential therapies for those diagnosed with this syndrome. Preliminary data suggests beneficial effects of B cell depletion therapy via anti-CD20 monoclonal antibodies (rituximab) in a subset of CFS patients [17], as well as low-dose naltrexone [18], but no treatment modality to date has consistently and conclusively been shown to be effective.

To provide some insight into the molecular processes that underlie CFS, we carried out a study on 15 patients diagnosed with CFS and 15 age, sex, and BMI matched controls **(Supplemental Table-2)**. CFS was formally diagnosed using the Fukuda Criteria **(Supplemental Table-4)**[19], Canadian Consensus Guidelines [20], and updated international consensus criteria, but excluding any related conditions such as major depressive disorder, Collagen-vascular diseases (CVD), neuromuscular diseases (NMD), and significant cardiac or pulmonary comorbidities **(Online Method)**. All participants were administered a moderate-intensity cardiopulmonary exercise test (CPET), with whole blood draws occurring immediately before the CPET and then 24, 48, and 72 hours post-CPET (**Fig. 1a**). In addition to the CPET, a clinical workup was performed on each participant, which included an EKG, standard metabolic panel blood test, height and weight, resting heart rate and blood pressure, and completion of a health assessment survey **(Online Method, Supplemental Table-3)**. Whole blood samples were cell sorted into B cells, Granulocytes, Monocytes, Natural Killer (NK) Cells, and T cells within 4 hours of collection and processed for RNA-seq. Differential Expression (DE) analyses showed no statistically significant (FDR<0.05) gene expression differences between any time points in either cases or controls for any of the cell types assessed. Relaxing the FDR threshold to <0.1 resulted in signatures detected in a few of the timepoints (CFS Bcell timepoints 1-2; CFS NK timepoints 1-4, 2-4, 3-4; CFS Mono timepoints 1-4; CFS Tempus timepoints 1-2, 2-4; Control Tempus timepoints 3-4). Furthermore, no statistically significant difference between time points was observed in data collected from the Modified Fatigue Impact Scale (MFIS) questionnaire, Karnofsky performance scores, or clinical workups, although patients did report increased physical fatigue (0-10 rating) between timepoint 1 and timepoints 2 and 3 **(Supplemental Table-3)**, consistent with previous findings that CPET does not appear to strongly effect molecular differences in CFS [21, 22] to a significant degree. These results suggest the need for a more intense exercise protocol and/or much larger sample sizes to assess whether post-exertional malaise may have a more modest impact on molecular states. Given the lack of a CPET-associated time signature, for all further analyses presented here, the time series data are treated as biological replicates.

**Fig. 1:**
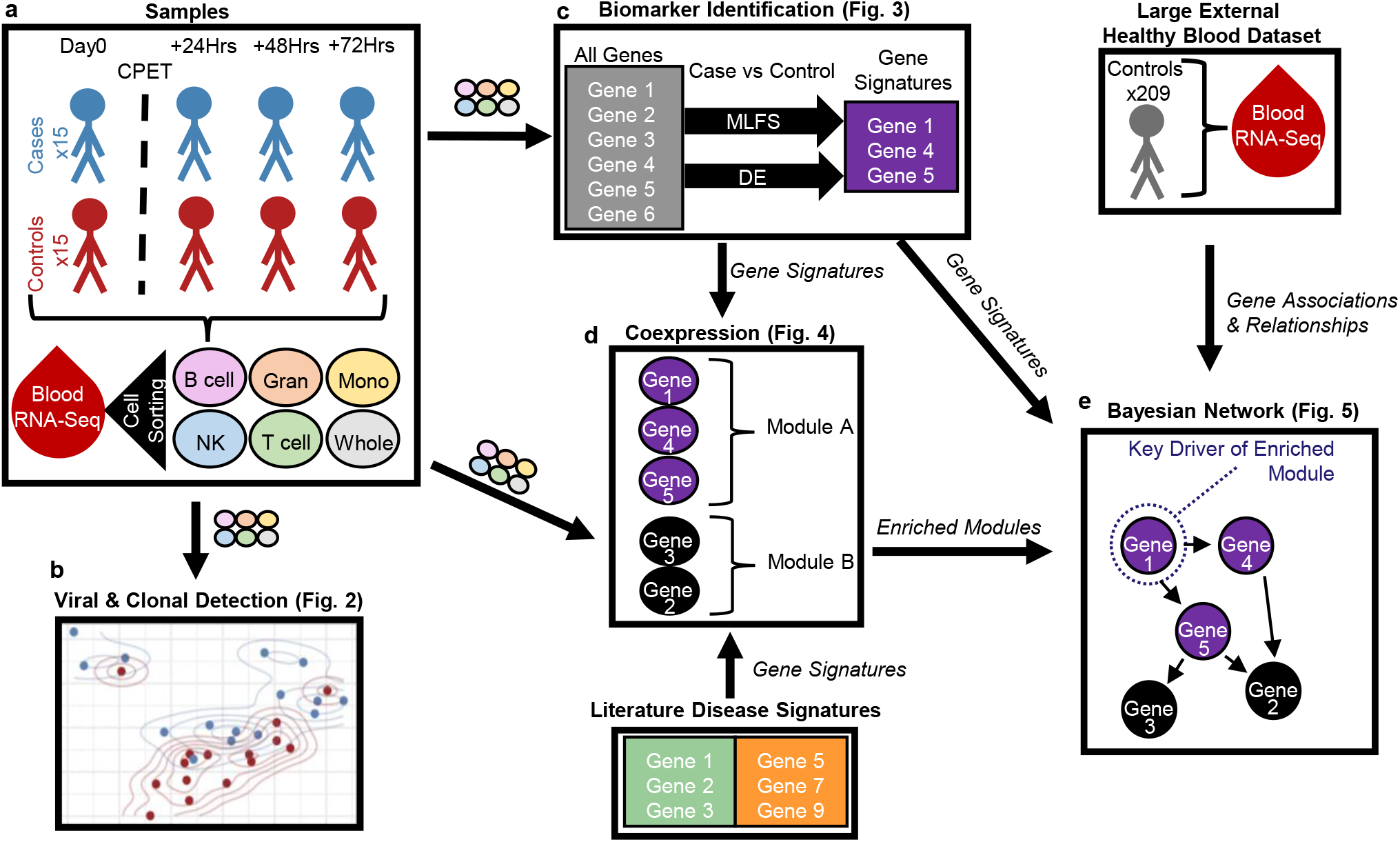
Study Analysis Workflow. A: RNA-seq read count data were generated on whole blood and FACs-sorted immune cell samples from CFS cases and controls. B: RNA-seq count data were passed through a viral-clonal detection pipeline. C: RNA-seq count data were passed through our MLFS and DE pipelines, generating predictive signatures of disease. D: Co-expression network construction organized genes into modules, which were annotated for biological pathways and other disease signatures. G: A union of modules with enrichment for CFS and CFS signatures were used with whole blood gene expression in an independent cohort to build a regulatory network where key drivers of disease were identified.

We applied a comprehensive RNA-seq data analysis pipeline (**Fig. 1b-e**.) to analyze whole blood, as well as specific immune cell types isolated from whole blood of all participants. The first part of our analysis involved passing the RNA-seq data through a viral/clonal detection pipeline further detailed in the methods section (**Fig. 1b**).To visualize virome-wide variance differences between cases and controls, we performed principal component analysis (PCA) on whole blood **(Fig. 2a)**. The first two PCs represented ∼30% of the total variance, revealing a separation between cases and controls across PC2 (∼10% of the total variance), suggesting differences in total viral loads between groups. To further characterize these differences, we tested for differences in the viral load distributions between cases and controls using a Wilcoxon rank (**Fig. 2b**) and Kolmogorov–Smirnov (**Fig. 2c**) tests. Both tests were significant at the p < 0.0001 threshold.

**Fig. 2:**
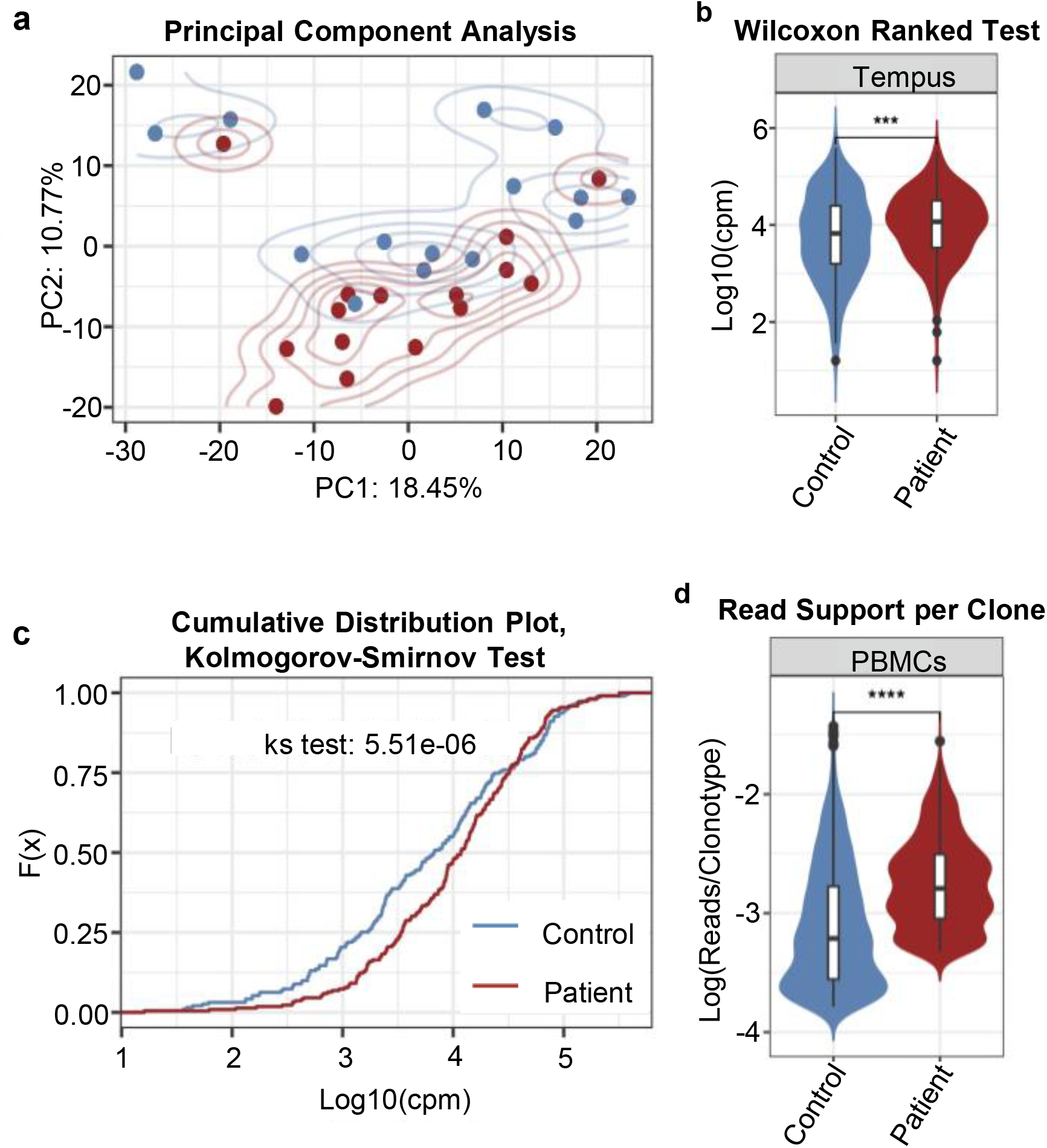
Viral and Clonal analysis detects dysregulation in CFS. A: Principal Component Analysis (PCA) of viral load estimated from the whole blood RNAseq data between patients and controls. B: Wilcoxon rank test of the viral mapping mean between patients and controls. C: Kolmogorov–Smirnov distribution test of the viral mapping mean between patients and controls. D: Clonal read support of T and B Cell clones between patients and controls.

Given the important roles T and B cells play in immune cell function, we investigated the clonality of each cell type by deconvolving the V[D]J genes using the MixCR algorithm [23]. We observed greater read support per clone or a less diverse population of T and B Cell clones in cases compared to controls (p < 0.0001) **(Fig. 2d)**, suggesting a dysregulation of these cell type populations in cases compared to controls.

To establish molecular signatures for CFS (**Fig. 1c, Supplemental Table-5**), we ran both DE (**Fig. 3b)** and Machine Learning Feature Selection (MLFS, methods) analyses (**Fig. 3a)**, leveraging both linear (DE) and non-linear (MLFS, random forest) approaches to identify gene expression features associated with case/control status. The non-linear MLFS strategy was used to cast the broadest net for identifying genes considered as significantly associated with case/control status (Information Gained Score FDR<0.05 in at least 1 timepoint per cell type), while the linear DE strategy was used to determine directionality of the gene expression changes between cases and controls for those genes identified by MLFS procedure (**Fig. 3c)**.

**Fig. 3:**
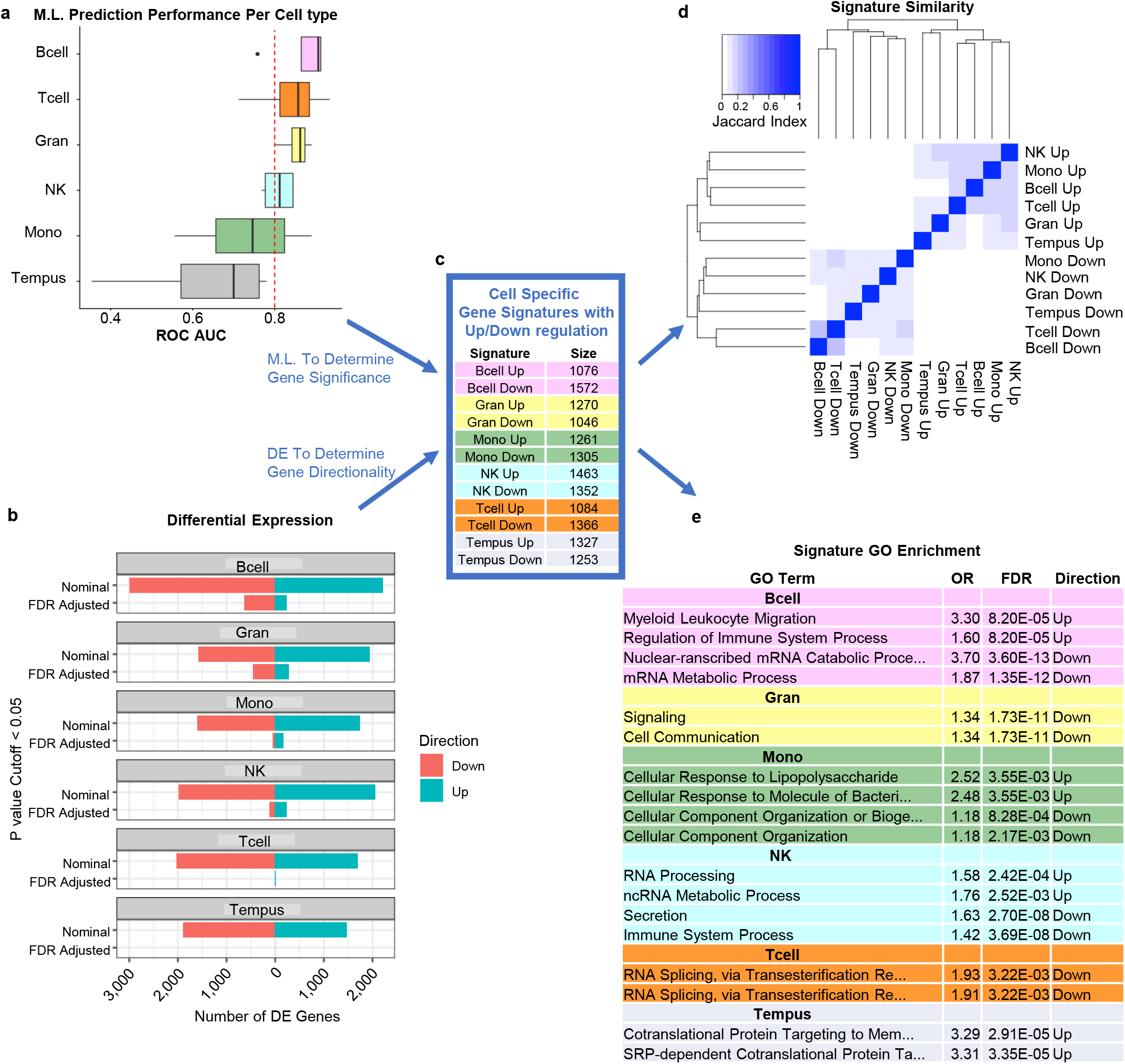
Machine Learning Feature Selection (MLFS) identifies predictive signatures of CFS. A: Classifier models built from CFS signatures show predictive ROC AUC performance on hold-out test sets across the different cell types (y-axis and color). B: DE analysis for CFS vs HC. X-axis represents the number of DE genes; bars are colored by direction of expression. Y-axis represents the different p value cutoff <0.05 of either Nominal or FDR adjusted p-value. C: CFS signatures were established using MLFS for significance and DE for expression directionality. Signatures are colored per cell type. D: Jaccard index showing signature similarity. E. Top GO enrichment table for signatures, colored per cell type.

Our MLFS strategy **(Supplemental Fig**.**1, Online Method)** resulted in 10 unique classifiers built per cell type, per timepoint, that were then tested against 10 unique hold-out test sets (**Fig. 3a)**. Classifiers built from B cell, T cell, Granulocyte, and NK cell signatures all had a mean receiver operating characteristic area under the curve (ROC AUC) > 0.80 (compared to an ROC AUC of 0.50 that would be expected by chance), with B cell signatures being the most predictive in terms of the mean ROC AUC (0.87 for B cells). Models built from whole blood had a ROC AUC = 0.63, suggesting that whole blood may be too noisy to detect a meaningful disease signature. The ability of our CFS classifiers to show predictability in this study is indicative of molecular differences between CFS cases and controls. These MLFS genes, along with their DE detected expression directionality, will be referred to as the CFS signatures for the remainder of this paper (**Fig. 3c)**.

To assess the similarity of the DE signatures across the different cell types, we computed the Jaccard index for all signature pairs (depicted as a heatmap in **Fig. 3d**). These results show that the signatures are largely unique across cell types, but that genes that are up (down) regulated within each cell type, cluster together across the cell types. **Fig. 3e** shows the top 2 gene ontology (GO) terms enriched in each direction of the cell-type specific CFS signatures, highlighting some of the biological processes (e.g. immune, metabolic and transcriptional dysregulation) that may be disrupted in CFS cases compared to controls. At the level of DE signatures, there is little overlap between the pathways that are the most significantly enriched across the cell types profiled.

To better characterize the subnetworks and pathways that may be disrupted in CFS, we constructed 6 co-expression networks to organize the gene expression data into coherent subnetworks of co-regulated genes (modules) for each of the 6 cell types (including whole blood) that were profiled in our study. While MLFS and DE aid in identifying the best features for distinguishing cases from controls, the co-expression network structure allows us to organize all of the gene expression traits into co-regulated groups of genes (modules) that place the individually identified MLFS/DE features into a more biologically relevant context. From the co-expression network analysis, we identified 119 co-expression modules spanning the 6 co-expression networks across all cell types (**Fig 1d, Supplemental Table-6**). Interestingly, 56 of the 119 modules were significantly enriched for the CFS signatures (**Fig 4a**).

**Fig. 4:**
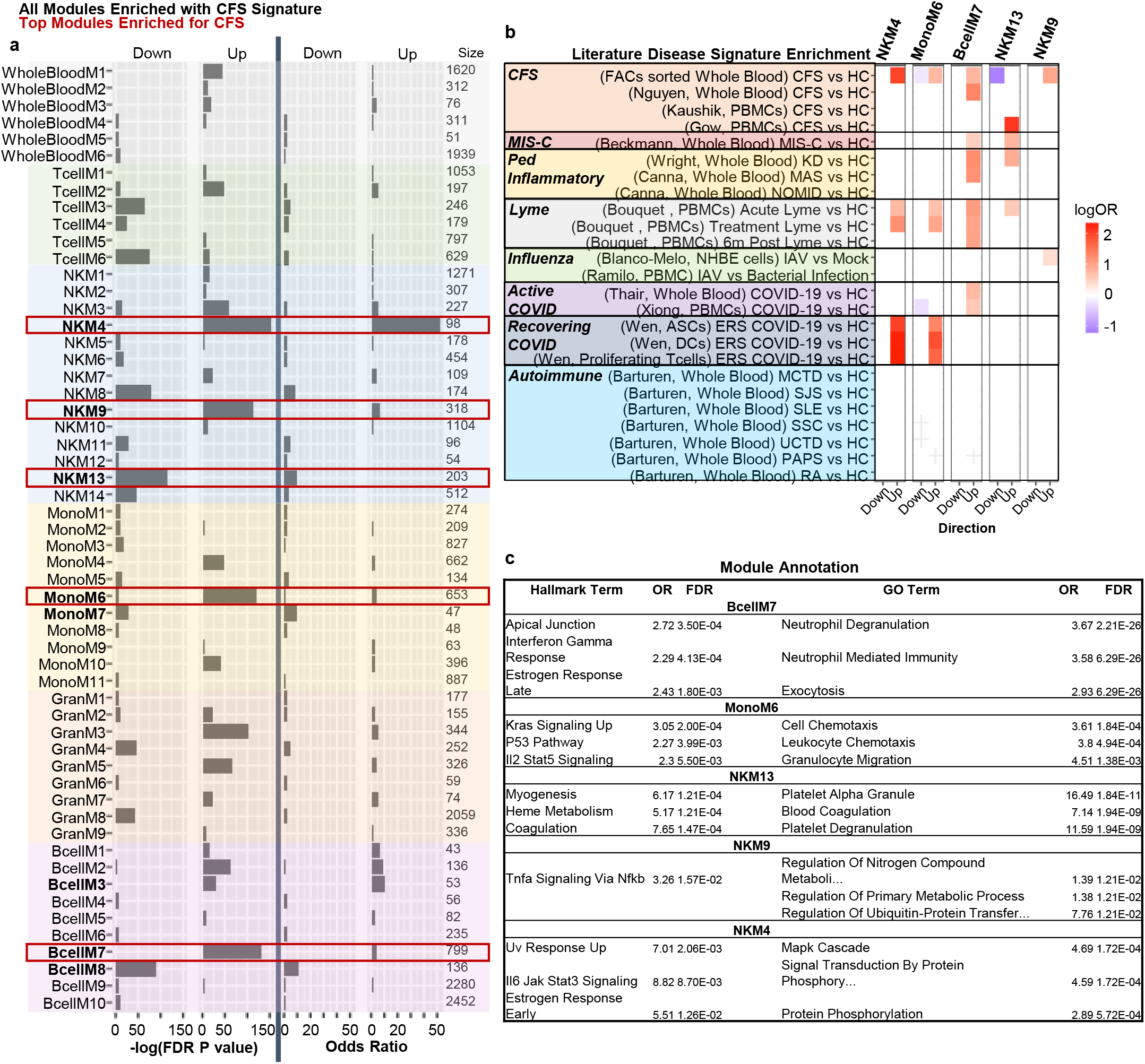
Co-expression network analysis identifies modules of genes dysregulated in CFS and other disease signatures. A: Table of all co-expression modules significantly associated with CFS signature, colored per cell type. X-axis represents either -log(FDR pvalue) or Odds Ratio (OR) of enrichment for CFS signature. The most significantly associated modules are highlighted with red boxes. B: Enrichment heatmap of top CFS modules and literature signatures from other disease. Y-axis represents disease signatures and are colored and grouped by the category of disease the signature falls into. C: Top 3 functional annotations of top CFS modules. This figure only shows those top modules with significant functional annotations.

We examined the top 5 modules enriched for the CFS signature as ranked by FDR adjusted p value: 1) Module 4 (M4) identified in the NK cell network (denoted as NKM4; FDR=1.39e-66, logFC= 1.71); 2) BcellM7 (FDR=2.15e-57, logFC=0.675); 3) MonoM6 (FDR=2.33e-53, logFC=0.658); 4) NKM13 (FDR=2.88e-50, logFC=1.002); 5) NKM9 (FDR=3.10e-50, logFC=0.826), for enrichment of published signatures (**Online Methods, Supplemental Table-7, 13**) for other fatiguing illnesses or diseases involving a strong inflammatory component such as Multisystem Inflammatory Syndrome in Children (MIS-C), Kawasaki Disease (KD), Macrophage Activation Syndrome (MAS), Neonatal Onset multisystem inflammatory (NOMID), Lyme disease, active Influenza (IAV), active COVID-19, early recovery stage after COVID-19, Mixed Connective Tissue Disease (MCTD), Sjögren’s Syndrome (SJS), Systemic Lupus Erythematosus (SLE), Systemic Sclerosis (SSC), Undifferentiated Connective Tissue Disease (UCTD), Primary Antiphospholipid Syndrome (PAPS) and Rheumatoid Arthritis (RA). The top CFS modules are enriched for many of these external disease signatures, such as MIS-C, Lyme disease, and COVID-19, but notably are not enriched for any of the autoimmune signatures (**Fig. 4b**). Similarities between CFS, COVID-19, and Chronic Lyme Disease have been suggested at a clinical level [3, 24, 25] and our data further suggests a shared molecular etiology among these diseases.

To further characterize these CFS modules, we searched for biological processes and pathways that were also enriched in them (**Fig. 4c, Supplemental Tables 8-10**). NKM4 was the module most strongly associated with CFS (logOR=1.71, FDR=1.39e-66) and was also enriched for the “recovering COVID-19” signature (Wen Proliferating T-cells, logOR=2.02, FDR= 1.07E-15), highlighting a molecular link that may help explain why so many recovered COVID-19 patients seem to experience CFS-like symptoms [2]. The top biological pathway associated with NKM4 was MAPK Cascade (logFC=4.691, FDR=1.72e-4), consistent with *in vitro* findings of MAPK dysregulation in NK cells of CFS [26] and COVID-19 [27]. Lyme disease signatures were enriched for the greatest number of top CFS modules, with 4 of the top 5 CFS-enriched modules also enriched for the chronic Lyme disease signature, suggesting a potential molecular link between CFS and chronic Lyme disease as has been previously clinically described [28]. Top biological pathways associated with these 4 modules include MAPK Cascade (logFC=4.691, NKM4 FDR=1.72e-4), KRAS Signaling Up (MonoM6 logFC=3.049, FDR=2.00e-4), Neutrophil Degranulation (BcellM7 logFC=3.668, FDR=2.21e-26), and Platelet Alpha Granule (NKM13 logFC=16.495, FDR=1.84e-11). These annotated module similarities appear to correlate with *in vivo* findings of MAPK dysregulation in NK cells of Lyme disease [29] and CFS [26], coagulation dysregulation in Lyme disease [30] and CFS [31], as well as neutrophil dysregulation in Lyme disease [32] and CFS [33] patients. MIS-C and Kawasaki Disease appear to share common molecular processes with CFS through the BcellM7 and NKM13 modules. Top biological pathways associated with these modules include Neutrophil Degranulation (BcellM7 logFC=3.668, FDR=2.21e-26) and Platelet Alpha Granule (NKM13 logFC=16.495, FDR=1.84e-11). These annotated module similarities align with findings of neutrophil dysregulation in Kawasaki Disease [34], MIS-C [35], and CFS [33] along with coagulation dysregulation in Kawasaki Disease [36], MIS-C [37], and CFS [31]. **Supplemental Table-7** shows the enrichment of all disease signatures across all of the CFS enriched modules.

While co-expression networks well organize the expression data into modules of coregulated genes, such networks to not predict regulatory relationships among the genes. To explore more dominant regulatory relationships in the top CFS co-expression modules, we constructed a probabilistic causal Bayesian network (BN) to aid in the identification of master regulators, or key driver genes (KDs), for each of the top CFS-associated modules (**Fig. 1e, Supplemental Table-1**2). Given our small sample size and to achieve adequate power in constructing a reliable BN model, we chose to use gene expression data from a large external gene expression dataset (Mount Sinai Crohn’s and Colitis Registry, MSCCR) consisting of 209 healthy individuals from which whole blood samples had been collected. We hypothesized that while gene regulatory relationships may be modulated at different levels in health individuals and those with CFS or the matched controls, the relationships themselves would be largely conserved as has been shown for other diseases [38-40]. We constructed the BN from a CFS-centered set of genes, including genes from the CFS and related disease signatures and co-expression modules significantly enriched for these signatures (13,332 genes in total) (**Fig. 1e, Supplemental Table-11**).

KDs for the 5 top CFS-associated modules were identified from our BN using Key Driver analysis (KDA)[39-42]. The KD analysis predicted 904 KDs within the network, where changes in any of the KDs are predicted to change the molecular states of genes enriched in one or more of the CFS-associated modules. **Fig. 5b-c** highlights KD properties of those KDs that were both global KDs as well as local KDs for CFS modules / signatures. Here we highlight the 11 KDs that were predicted to modulate at least 3 CFS modules/ signatures **(Fig. 5b)**: MXD1, STX3, DYSF, LYN, MLL2, NCOA2, PTPRE, REPS2, RP11-701P16.2, TECPR2, and TUBB1. These KD genes have been shown to be associated with other diseases such as Familial hemophagocytic lymphohistiocytosis 5 (FHL5) [43], chronic myeloid leukemia [44], Dysferlinopathy and inflammatory myopathy [45], Lupus [46], B cell lymphoma [47] as well as immunological aspects of Kabuki syndrome [48], Kaposi’s sarcoma-associated herpesvirus [49], modulator of macrophage activation and inflammatory diseases [50], prostate cancer [51], spastic paraparesis [52], and abnormal platelet physiology [53].

**Fig. 5:**
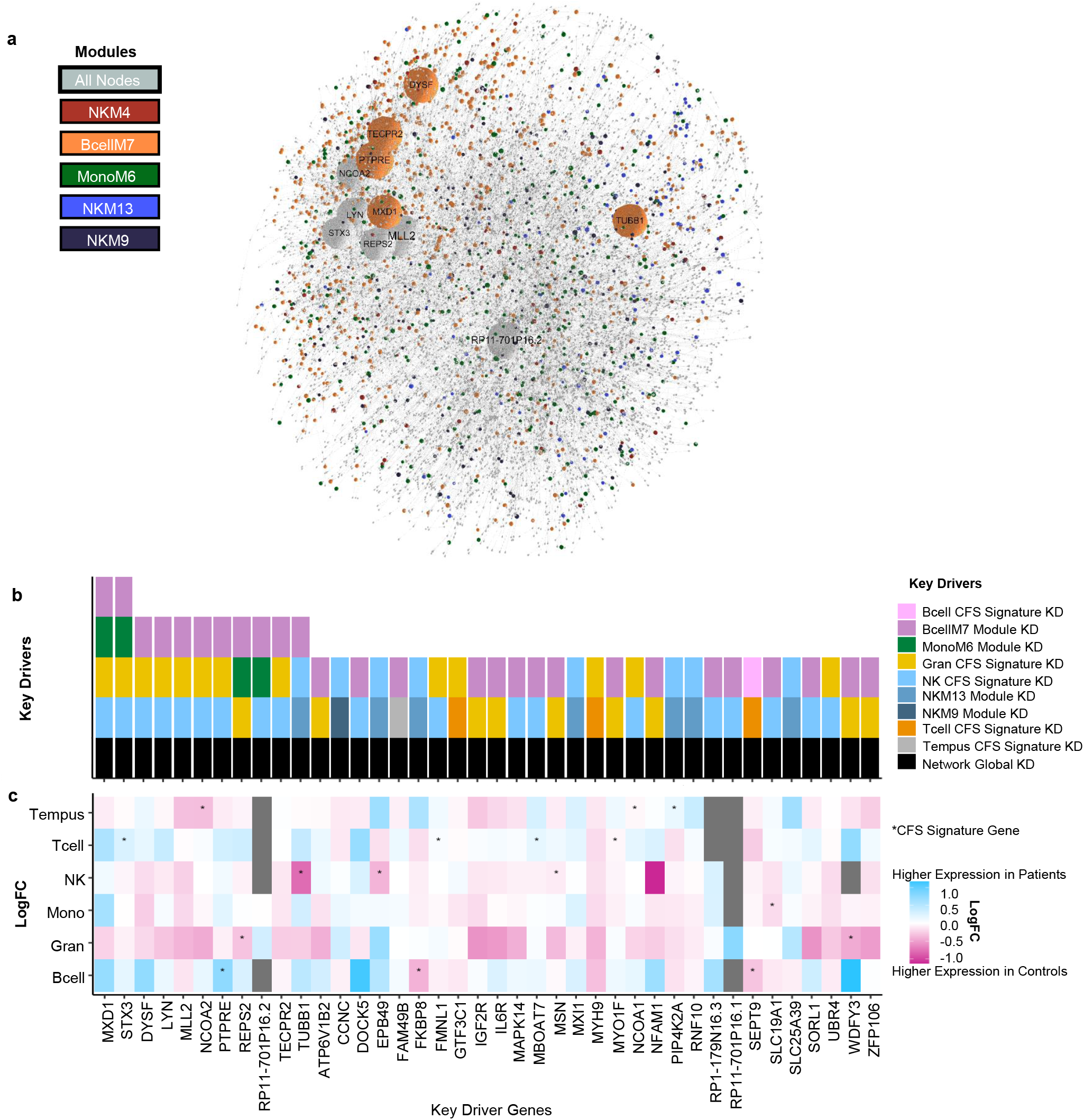
Bayesian Network and Key Driver Analysis identifies regulators of CFS. A: A 2D representation of the Bayesian network. B and C illustrate key drivers of the network. B: shows the frequency in which a gene is considered a KD. C: shows the DE logFC of the KD gene in the cell specific signatures.

To provide experimental support for the KD genes identified from the network, we computed the probability of being loss-of-function intolerant (pLI)[54] across all of the KD genes and compared those scores to non-KDs. The pLI results showed that our KDs predicted to modulate CFS-associated network modules have an increased pLI when compared to the non-KD genes (median pLI for non-KDs = 0.02, median pLI for KDs = 0.68, one-sided Wilcoxon test p value = 1.80e-48), supporting that the KDs are biologically important to the network **(Supplemental Fig. 2, Online Method)**. The combination of high pLI scores and known dysregulation leading to disease states support these CFS KDs as master regulators of vital biological processes associated with CFS. Many of these KDs have been previously shown to be KDs of immunological dysregulation diseases but to the best of our knowledge, have not previously been associated with CFS.

In conclusion, we present an unbiased data driven, network-based approach that identified molecular signatures of CFS and implicates a number of highly coherent co-expression modules to CFS. For the top 5 modules, the complementary analysis shown here point to a common underlying biology that shares immune and metabolic dysregulation also present in other clinically similar diseases such as Lyme, MIS-C, Kawasaki, and recovering COVID-19. Moreover, the top KDs we identified as regulators of these CFS-associated modules are biased towards higher pLI scores, indicating that loss of function mutations in these genes cannot be well tolerated, confirming their critical importance to normal system function. These top KDs we identified for CFS offer interesting points of therapeutic intervention to explore, with the most promising being MXD1, STX3, DYSF, LYN, MLL2, NCOA2, PTPRE, REPS2, RP11-701P16.2, TECPR2, and TUBB1. To help facilitate continued CFS community research we are also providing an interactive website containing the signatures, modules, KDs, and Bayesian network which can be found here: https://irenefp.github.io/bcellm7.html

## Data Availability

All data referred to in this manuscript will be included in this submission. An interactive website containing the signatures, co-expression modules, key drivers, and Bayesian network can be found here: https://irenefp.github.io/bcellm7.html

## Supplementary Tables

1. Covid-CFSCommonSymptoms
2. StudySummary
3. ClinicalWorkupQuestionnaire
4. Fukuda
5. Signautures
6. Modules
7. ModuleDiseaseEnrichment
8. ModuleGOAnnotations
9. ModuleMSigHallmarkAnnotations
10. ModuleMSigC7Annotations
11. BayesianNetwork
12. KeyDrivers
13. SignatureRefenences

## ACKNOWLEDGEMENTS

ZHG acknowledges funding from grant UL1 TR001863/TR/NCATS NIH HHS/United States.

## Methods

### Cohort Generation

The study consists of 15 patients and 15 age, sex, race, and BMI matched control participants who were recruited at the Mount Sinai Hospital. Informed consent was obtained from either the participant or their legally authorized representative, and all study-related activities were conducted under the approval and oversight of Mount Sinai’s Institutional Review Board (IRB). Experiment sequence: (1) Clinical evaluation and enrollment; (2) Symptom questionnaire **(Supplemental Tables 3-4)**; (3) Blood draw prior to Cardiopulmonary Exercise Testing (CPET); (4) Standardized CPET and Symptom questionnaire immediately after CPET; (5) Symptom questionnaire and blood draws 24, 48, and 72 hours post-CPET.

Inclusion and Exclusion Criteria:

#### Inclusion

- Patients met diagnostic criteria according to:
- Fukuda et al. [19] AND
- The Canadian Consensus Guidelines [20] AND
- The updated international consensus criteria [55]
- 18-57 years of age
- Willing to undergo phlebotomies, one at baseline, one pre-CPET, one 24 hours post CPET, one 48 hours post CPET and one 72 hours post CPET
- Willing and physically able to participate in cardiopulmonary exercise testing, i.e., exclusion of any absolute contraindications

#### Exclusion

- Diagnosis of major depressive disorder prior to diagnosis of ME/CFS
- Significant cardiac or pulmonary comorbidities
- Collagen-vascular diseases (CVD) and neuromuscular diseases (NMD)
- Anemia with hemoglobin levels of <10mg/dl
- Active malignancy (but not history thereof)
- Patients unwilling to undergo a wash-out of all immunomodulatory medications for a two-week period prior to cardiopulmonary exercise testing
- Pregnancy
- Any active acute infectious or acute inflammatory state (i.e., viral upper respiratory tract infection, cellulitis, urinary tract infection, sun burn, gastroenteritis, acute diarrheal illness etc.), any current or recent systemic antibiotic therapy (within one month prior to cardiopulmonary exercise testing)
- Inability to perform basic cycle or treadmill ergometer cardiopulmonary exercise testing
- Presence of any absolute contraindications for cardiopulmonary exercise testing:
- Acute myocardial infarction
- Unstable angina
- Uncontrolled arrhythmias causing symptoms or hemodynamic compromise
- Syncope
- Active endocarditis
- Acute myocarditis or pericarditis
- Symptomatic severe aortic stenosis
- Uncontrolled heart failure
- Acute pulmonary embolus or pulmonary infarction
- Thrombosis of lower extremities
- Suspected dissecting aneurysm
- Uncontrolled asthma
- Pulmonary edema
- Room air hypoxemia
- Respiratory failure
- Mental impairment leading to inability to cooperate with testing
- Presence of relative contraindications to cardiopulmonary exercise testing (based on physician assessment at time of testing):
- Severe untreated arterial hypertension
- High degree AV blockade
- Hypertrophic cardiomyopathy
- Significant pulmonary hypertension

Control participants were matched by race, age, sex and BMI. For the matching process, the variables age and BMI were categorized and matched by category rather than continuous value. The age categories were: 18-22, 23-27, 28-32, 33-37, 38-42, 43-47, 48-52 and 53-57. The BMI categories were: <19, 19-22, 23-26, 27-30, 31-34, 35-38, 39-42, 43-46 and >46 **(Supplemental Table-2)**. A standardized Control Subject Telephone Screening (CSTS) form was used to determine eligibility of control participants based on matching needs for the study.

### CPET

Patients were asked to refrain from physical exercise greater than 5 min of walking or the equivalent thereof for 2 days before and 3 days after the CPET (pilot phase) or until the post-exercise time point is reached (either 24, 48 or 72 hours post exercise; main phase). All CPET testing was performed between 8am and 11am. Visual analogue scale (VAS) symptom assessments via modified Fatigue Impact Scale (MFIS) were performed during enrollment, immediately prior, immediately after, as well as 24hrs, 48 hrs and 72hrs after exercise (see Modified Fatigue Impact Scale (MFIS) Sheet in Appendix). Also, on a simplified VAS, global parameters of (1) mental fatigue, (2) physical fatigue and (3) overall body pain were assessed at enrollment, immediately prior to CPET, every 5 minutes during the CPET, immediately after CPET, as well as 24hrs, 48hrs, and 72hrs after CPET (see simplified VAS assessment sheet in Appendix). Patients underwent cycle or treadmill ergometry, with primary option being cycle ergometry due to better signal quality, but with treadmill ergometry being an alternative for patients unable to perform bicycle ergometry. Patients during the first 5 minutes were asked to gradually increase their work rate until reaching 70% of age-predicted maximal heart rate [56], at which point this target heart rate was maintained. Ratings of perceived exertion (RPE) were obtained on a VAS from 1-10 every 5 minutes while undergoing testing (5, 10, 15 and 20min timepoints). Blood pressure measurements and lactic acid measurements (Lactate Pro Portable Analyzer (DKD, Japan) were performed every 5 minutes while exercising. The maximum duration of exercise testing was 25 minutes.

### Peripheral blood processing

Peripheral blood was collected into Na Heparin vacutainers for immune cell isolation and Tempus RNA vacutainer for total cellular RNA isolation. All blood were processed within 3 hours of phlebotomy. Blood collected in Na Heparin tubes was subjected to a modified Ficoll gradient centrifugation to isolate both peripheral mononuclear cells (PBMCs) and granulocytes. Briefly, following the Ficoll centrifugation, two different cellular fractions were collected, the unique buffy coat layer (PBMCs) and granulocyte on top of the red blood cell layer [57]. PBMCs were washed twice to remove the platelet and used for subsequent four immune cell subset isolations, T cell, B cell, NK cells and monocytes using the Stem cell EasySep system (STEMCELL Technologies Inc.) following the manufacturer’s recommendations. The number, viability and the purity of the isolated immune cell subsets were determined by cell counters and flow cytometry as per standard procedures. Only the cells meeting the Quality Control measures were then lysed in RLT buffer with 1% beta-mercapto-ethanol and kept frozen at −80C until commencement of batch sample RNA isolation. The resulting immune cell lysate fractions were: Whole blood cells (from Tempus vacutainer), Granulocytes, Monocytes, B cells, T cells, Natural Killer cells. RNA was extracted using RNeasy kits (Qiagen) and treated with RNAse free DNase-I per manufacturer’s instruction (Qiagen) to remove the any contaminated DNA. Quality and Quantity of the RNA were analyzed by Bioanalyzer (Agilent Technologies Inc.) and Nanodrop (ThermoFisher SCIENTIFIC). RNAs were kept frozen stored at −80C until RNAseq library synthesis.

### RNA-Seq Library preparation

About one microgram of total RNA was used for the preparation of the seq library using TruSeq mRNA Seq Kit supplied by Illumina (Cat # 1 FC-122-1001). The protocol followed was as per manufacturer’s instruction. Briefly, mRNA was isolated from total RNA using oligo dT on magnetic beads. The mRNA was then be fragmented in the presence of divalent cations at 940C. The fragmented RNA was converted into double stranded cDNA. After polishing the ends of the cDNA, adenine base was added at the 3’ ends following which Illumina supplied specific adaptors were ligated. The adaptor ligated DNA was amplified by 15 cycle PCR. The PCR DNA was purified on Ampure beads (Beckman Coulter, part# 63882) to get the final seq library ready for sequencing. The insert size and DNA concentration of the seq library was determined on Agilent Bioanalyzer. Each RNA seq library was layered on one of the eight lanes of the Illumina flow cell at appropriate concentration and bridge amplified to get around 350 million raw reads. The cDNA reads on the flow cell will then sequenced on HiSeq2500 using 100 bp single end recipe. Five barcoded samples were pooled to sequence in one lane.

### Sequencing

The RNA-Seq libraries were sequenced on the Illumina HiSeq 2500 platform using 100 bp single end protocol following manufacture’s recommended procedure. Base calling from Images and fluorescence intensities of the reads were done in situ on the HiSeq 2500 computer using Illumina software. Various QC parameters such as intensities of individual bases, visual and graphic focus quality of the images were monitored periodically to assess the quality of ongoing run. Sequence quality was monitored in terms of colored graphic representation of Q30 values (which is a measure of errors per thousand base), and error rates at 35 and 75 cycles of sequencing were observed to assess the quality of ongoing run. The seq data generated was simultaneously transferred (in real time manner) to high performance computer cluster and processed for Picard based QC analysis. The Seq data passing all QC parameters was used for further analysis.

The raw RNA-seq reads were converted to featureCounts after being aligned to HG19 reference using STAR and the RAPiD [58]pipeline (v2.0.0). Reads not mapped to human HG19 were later analyzed with the Viral Load pipeline described below in these methods. FeatureCounts data were separated into their respective cell types and converted to countspermillion (cpm) with edgeR [59]and filtered so that only genes with >1 cpm in 10% of the data were kept (Bcell=13,262 genes, Granulocytes=10,612 genes, Monocyctes=11,947 genes, NK cells=13,391 genes, Tcells=13,347 genes, Whole Blood=13,415 genes). Gene counts were normalized using the limma [60] functions calcNormFactors(method=“TMM”) and voom() per cell type.

We used principal component analysis prcomp() [61] and variancePartition [62] package to explore and visualize the gene count expression variance from covariates. Due to the small samples size and the age, sex, BMI, matched study design, no covariates were adjusted for during normalization.

### Differential Expression

The limma package [60] was used to perform differential expression (DE) analysis performed on the RNA-seq normalized counts data per cell type. The function duplicateCorrelation() was used to mitigate multiple measurements from the separate timepoints, followed by lmfit(), contrasts.fit(), and eBayes(). DE analysis revealed no significant (FDR<0.05) genes were found to be differentially expressed among any timepoint pairs within CFS and control status. Due to this result, timepoint data was treated as biological replicates for all further analysis. DE analysis did reveal differentially expressed genes between CFS and control status within each cell type.

### Machine Learning Pipeline

We used machine learning classifiers to unbiasedly reduce dimensionality in the RNA-seq normalized counts data to genes most informative to accurately classify CFS and control samples, known as machine learning feature selection (MLFS) [63] **(Supplemental Fig. 1, Supplemental Table-5)**. This pipeline was performed on each cell type and type point separately to reduce data leakage in the training/testing splits. MLFS was performed in 5-Cross Test Folds, where the data were randomly split into training (24 samples) and testing (6 samples), and stratified by class as to not generate imbalance [64]. The training data were further split in a 5-fold cross validation resulting in trainingII and validation. Random forest classifiers (python sklearn package [65, 66]) were fit on each of the 5 trainingII splits, where the number of trees in the forest is equal to the number of gene features in the data set, resulting in large random forests that gives most, if not all, genes a feature importance (information gained) score [67]. A Bonferroni adjusted p-value was fit to the information gained scores and genes with adjusted pvalue <0.05 were considered significant [68]. This was done for each of the 5 trainingII subsets, results in 5 lists of significant features per cell type, per timepoint. The union of the 5 lists of significant features were pushed to the next step where a new random forest classifier was then fit on the entire training data samples, but with only the significant features found in the previous step. This new classifier was then tested against the testing data and classifier performance was recorded **(Fig. 3a)**. The final result is 5 distinct classifier models, built from each of the 5-Cross Test Folds, along with feature importance ranked lists of all gene features in the dataset. By setting up our machine learning pipeline this way, we used classifiers to find gene features that consistently separate patients from controls, regardless of random effects of splitting the dataset. This was done independently for each timepoint and each cell type so that at no time could an individual be in both the training and testing during the same run through.

### Weighted Gene Coexpression Network Analysis (WGCNA)

WGCNA co-expression networks were built upon each of the different cell types independently, treating biological replicates as independent samples, resulting in 6 co-expression networks **(Supplemental Table-6)** [69]. Network building parameters included softpower thresholds of 14 in the adjacency() function, dynamic tree module cutting, and no samples were excluded from sample trees. To identify modules enriched for CFS disease signal, we calculated enrichment statistics using Fisher’s Exact Test (fisher.test() [61]), and corrected for multi-testing following Benjamini-Hochberg procedure [70] (p.adjust () [61])of the MLFS signature genes on each of the co-expression networks **(Supplemental Table-7)**.

### Pathway Enrichment Analyses

Pathway enrichment for GO [71, 72], C7, and Hallmark [73] pathways (N=10,192, 4872, and 50 pathways, respectively) was performed on the MLFS CFS signatures and all co-expression modules **(Supplemental Tables 8-10)**. Enrichment was performed separately for each cell type modules and signatures as each cell type had different dataset background (Bcell=13,262 genes, Granulocytes=10,612 genes, Monocyctes=11,947 genes, NK cells=13,391 genes, Tcells=13,347 genes, Whole Blood=13,415 genes) and Benjamini-Hochberg multiple testing correction was performed for each cell type separately as well using p.adjust() function in R [61]. GO enrichment was performed using the R packages goseq [74], topGO [75] and org.Hs.eg.db [76] while C7 and Hallmark enrichment was performed using the R packages HTSanalyzeR [77], GSEABase [78], and GAGE [79].

### B cell and T cell Clonal Detection / Viral Load

To recover information on the clonal composition of T and B-cells, we used the software MIXCR with default parameters over RNA-seq fastq files [23]. Briefly, the tool performs an alignments of sequencing reads to reference V, D, J and C genes of T- or B-cell receptors and filters sequences based on read quality. Next, the assembly of clonotypes is performed using the previous alignments including calibration and correction based on upstream and downstream regions of the target genes in order to extract specific CDR3 regions. Finally, the alignments, clonotypes, and molecular identifiers are exported into a tab delimited text file. The outputs are carefully reviewed and a threshold of at least 10 reads per clonotype is set for further downstream analysis. Visualization and figures are done with ggplot [80] and cowplot [81]packages available in R statistical software [61].

To identify reads that correspond to possible viral species, we used the non-human or non-mapping reads from the main alignment used in this study. First, these reads were assembled into contigs using the aligner inchworm from Trinity Suite [82] stopping before the chrysalis step. Next, we filtered out the alignment for contigs smaller than 200 base pairs. Also, we further refine the contigs by consolidating contig clusters with a similarity of 95% or above by using the software cd-hit [83] with the parameters “-c 0.95 -n 10 -T 2 -p 1 -g 1”. Next, we perform alignment with diamond suite [84] against the NCBI viral genome assemblies with the following parameters: “-f 6 qseqid sseqid pident length mismatch gapopen qstart qend sstart send evalue bitscore qframe salltitles -k 1”. Finally, the matching alignments were combined with the identifiers per contig in the R statistical software [61] and used in downstream analysis with the limma [60]/edgeR [59, 85] linear modeling and ggplot2 [80]/cowplot [81]packages.

### Bayesian Causal Network

209 healthy whole blood RNA-seq samples (residual counts matrix after adjusting for demographics of age, gender and genetic PC’s, and technical covariates RIN, processing batch and ribosomal RNA rate) were acquired from the Mount Sinai Crohn’s and Colitis Registry [86] (MSCCR) to be used in the Bayesian Network **(Supplemental Table-11)**. The Bayesian Network was built using RIMBANET [87-90]. Seeding gene list was generated with the union of MLFS signature genes and genes found in all co-expression modules enriched for MLFS signatures, resulting in a seeding gene list of 13,332 genes. Reducing the network search space to the seeding gene list is important for increasing the the fit of the network. MSCCR healthy RNA-seq blood sample expression, with their associated eQTLs were used as priors to build the network [86].

### Interactive Network Visualization

For intuitive visual exploration of the generated dense and complex networks, we have developed a web-based interactive CFS network exploration portal, which builds from our previous work on interactive network visualization [91, 92]. The portal provides a customized exploration environment for the network and other associated meta-data. Briefly, we have implemented the 3D capability by building with Three.js, which is a cross-browser JavaScript library that incorporates the WebGL (Web Graphics Library) API. We have calculated the network layout using the 3d-force-graph, which is a library for Three.js. In addition, we have utilized client-side Javascript libraries d3.js and jQuery in real time. For the styling of web interface elements, we have primarily utilized custom HTML and CSS. The portal utilizes only standard libraries and does not require the use of any additional plug-ins, which enables it to run on all modern web browsers. Furthermore, users can easily define filters on clinical variables (modules, key drivers). The portal is an open-access freely available resource for the dissemination of all our network analysis results to the scientific community. The portal can be accessed at: https://irenefp.github.io/bcellm7.html.

### Key Driver Analysis

Key Driver Analysis was performed using KDA [93] **(Supplemental Table-12)**. For local key drivers, this package defines a background sub-network by looking for a neighborhood K-step away from each node in the target gene list in the network. Stemming from each node in this sub-network, it assesses the enrichment in its k-step (k varies from 1 to K) downstream neighborhood for the target gene list. In this analysis, we used K=6. Target gene lists used for KDA included MLFS signature genes and enriched co-expression module genes. This package defines global drivers or hub genes as genes with number of neighbors exceeding that of the mean of gene neighbors + 2 standard deviations of gene neighbors of all genes in the network.

### pLI

We mapped probability of being LoF intolerant (pLI) score [54] to all genes in the Bayesian Network **(Supplemental Fig. 2)**. To test for differences between KD and non KD genes, we used a one-sided Wilcoxon test.

### Curated External Disease Signatures

Curated external disease signatures from the literature were used to gain further insight into shared disease etiology in our co-expression network. Further signature information can be found in **Supplemental Table-13**. The following signatures were used in our analyses:

- (Nguyen, Whole Blood) CFS vs HC [94]
- (Kaushik, PBMCs) CFS vs HC [95]
- (Gow, PBMCs) CFS vs HC [96]
- (Beckmann, Whole Blood) MIS-C vs HC [97]
- (Wright, Whole Blood) KD vs HC[98]
- (Canna, Whole Blood) MAS vs HC, (Canna, Whole Blood) NOMID vs HC [99]
- Lyme vs HC, (Bouquet, PBMCs) Treatment Lyme vs HC, (Bouquet, PBMCs) 6m Post Lyme vs HC [100]
- (Blanco-Melo, NHBE cells) IAV vs Mock [101]
- (Ramilo, PBMC) IAV vs Bacterial Infection [102]
- (Thair, Whole Blood) COVID-19 vs HC [103]
- (Xiong, PBMCs) COVID-19 vs HC [104]
- (Wen, ASCs) ERS COVID-19 vs HC, (Wen, DCs) ERS COVID-19 vs HC, (Wen, Proliferating Tcells) ERS COVID-19 vs HC [105]
- (Barturen, Whole Blood) HC MCTD vs, (Barturen, Whole Blood) SJS vs HC, (Barturen, Whole Blood) SLE vs HC, (Barturen, Whole Blood) SSC vs HC, (Barturen, Whole Blood) UCTD vs HC, (Barturen, Whole Blood) PAPS vs HC, Barturen, Whole Blood) RA vs HC [106]

### Miscellaneous Statistical/Computational Analyses

R analysis in this paper were performed using R version 4.0.1. R package libraries include:

data.table (v1.13.0)

edgeR (v3.30.3)

limma (v3.44.3)

variancePartition (v1.19.4)

WGCNA (v1.69)

goseq (v1.40.0)

topGO (v2.40.0)

org.Hs.eg.db (v3.11.4)

HTSanalyzeR (v2.34.1)

GSEABase (v1.50.1)

GAGE (v2.38.3)

ggplot (v3.3.2)

cowplot (v1.1.0)

RIMBANET

KDA

Python analysis for the MLFS was performed using python version 3.7.3. Python packages include:

numpy (v1.16.4)

pandas (v0.24.2)

scikit-learn (v0.21.2)

---

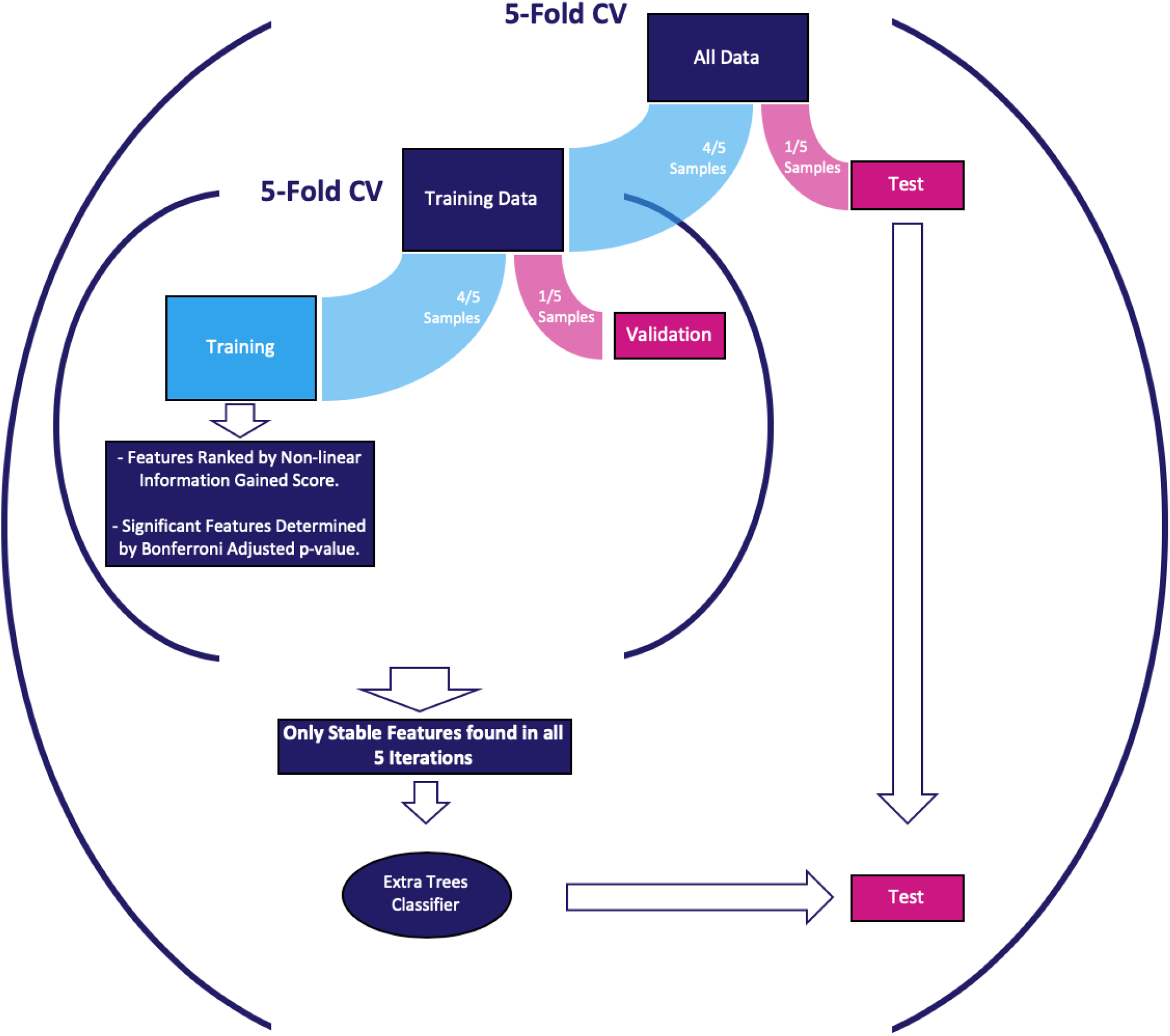

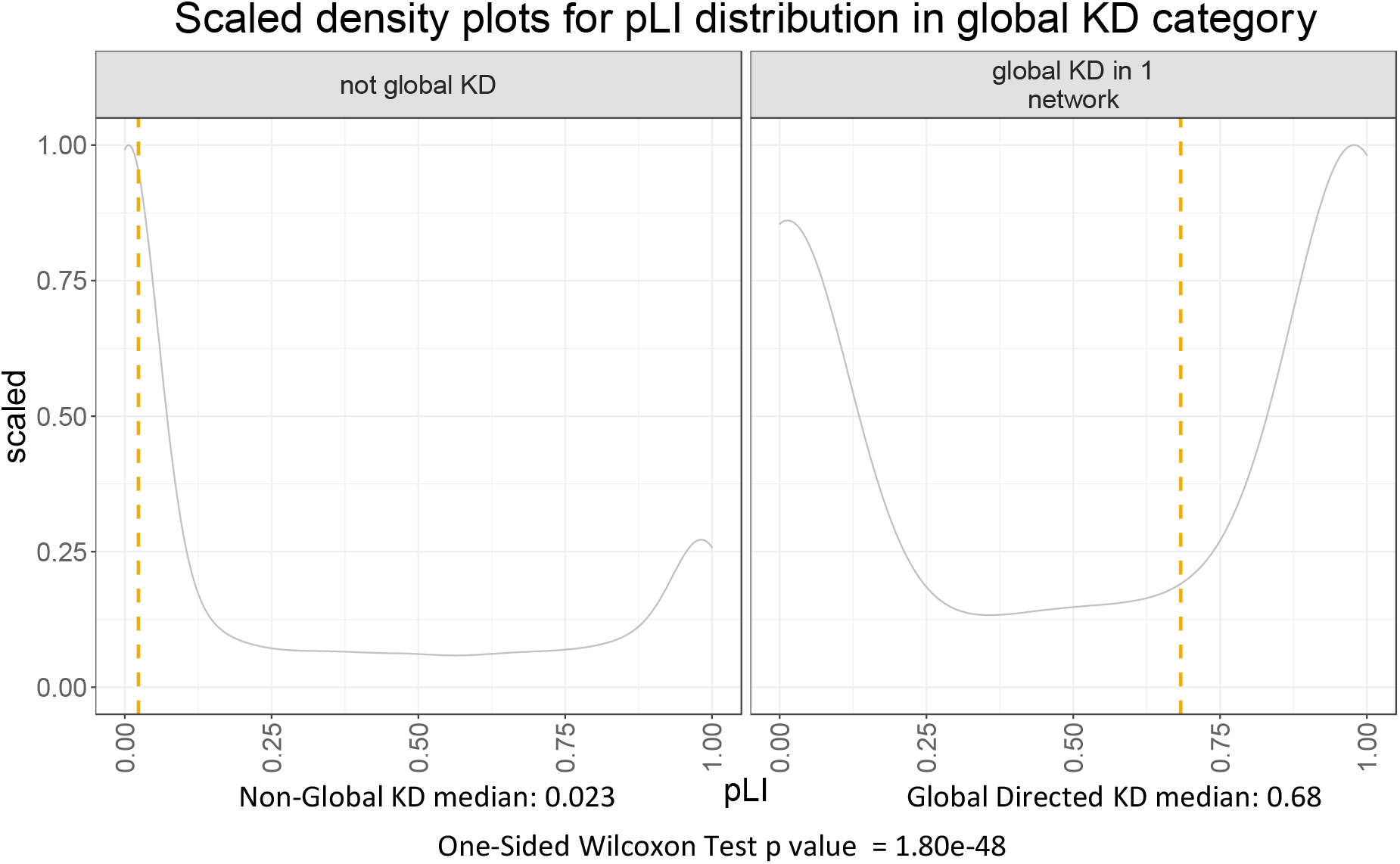

